# SNRI-driven recovery of impaired pupil dynamics in Progressive Supranuclear Palsy

**DOI:** 10.1101/2025.10.14.25337762

**Authors:** Molly Zeitzschel, Maxime Maheu, Tobias H. Donner, Christian K. E. Moll, Carsten Buhmann, Götz Thomalla, Tim U. Magnus, Günther U. Höglinger, Keno Hagena, Alessandro Gulberti, Monika Pötter-Nerger

## Abstract

**Background:** Progressive supranuclear palsy (PSP) is a tauopathy marked by early degeneration of brainstem nuclei such as the locus coeruleus (LC), a central player of the ascending reticular activating system (ARAS). ARAS dysfunction contributes to cognitive and arousal disturbances and is increasingly recognized as a therapeutic target.

**Methods:** 14 PSP patients (eight females, age 70.81 ± 6.32 years, disease duration 4.69 ± 3.24 years) were assessed before and after ≥4 weeks of SNRI (serotonin–norepinephrine reuptake inhibitor) treatment. Participants were assessed by non-invasive neurophysiological measures as pupillometry, performed at rest and during an auditory oddball paradigm, and clinically, using PSP-specific clinical scales and quantitative gait analysis. Pupil, cognitive, and gait parameters were compared with age-matched healthy controls.

**Results:** At baseline, pupil size, pupil dilation responses and surprise pupil responses were reduced in PSP patients compared to controls, accompanied by impaired executive functions, reduced phonemic verbal fluency and depressive symptoms in PSP patients. SNRI treatment selectively rescued certain impaired pupil metrics in PSP, leading to a significant improvement of the surprise pupil response and partial normalization of pupil fluctuations at rest. Pupil metric changes were associated with notable improvement in executive functions and quality of life at follow-up.

**Conclusion:** In summary, our findings reveal altered pupil-linked arousal regulation in PSP and its modulation by SNRI therapy, suggesting pupillometric indices as promising biomarkers of therapeutic response.

## Introduction

Progressive supranuclear palsy (PSP) is a neurodegenerative disease, pathologically defined by 4R-tau protein aggregation and clinically characterized by core clinical features as oculomotor dysfunction, postural and gait disturbance, parkinsonism, and cognitive deficits with symptom patterns varying across PSP subtypes.^1^ The disease is increasingly recognized as a network disorder with predominant involvement of brainstem and subcortical structures leading to cortico-brainstem and subcortico-cortical connectivity disruption.^2–4^

The early involvement of brainstem structures in PSP involves the ascending reticular activating system (ARAS),^2^ comprising a number of reticular nuclei and their widespread ascending projections to thalamus, hypothalamus, basal forebrain and cortex.^5^ The ARAS is supposed to regulate arousal and cognitive processes.^6^ ARAS-related functions, which are known to be impaired in PSP, include arousal, attention, executive functions and verbal fluency.^6–8^ A key component of the ARAS, the noradrenergic locus coeruleus system (LC), is particularly affected in PSP. Degeneration of the LC is an early feature that is evident across PSP subtypes,^3,9^ and its degree correlates with disease severity and cognitive impairment.^9,10^ Tau pathology extends further to the serotonergic dorsal raphe nuclei, and tracer uptake in these regions correlates with clinical disease severity.^2,11,12^

The ARAS has a central role in controlling pupil-linked arousal.^13,14^ Particularly, the activity of brainstem-cortex bidirectional projections is reflected in changes in pupil size.^13,15^ Animal studies^16–18^ and one MRI study in humans^19^ report a close correlation of LC activity with slow pupil fluctuations at rest,^14,16,17,19^ and stimulus-evoked pupil dilation responses.^17–20^ As a consequence, non-luminance-mediated changes in pupil size are widely used as a non-invasive index of LC activity.^14^ Furthermore, other nuclei within the ARAS, such as the serotonergic dorsal raphe system, are linked to pupil size changes.^17,21,22^

To date, involvement of the ARAS in PSP has been reported only indirectly through a limited number of neuropathological and neuroimaging studies, and it has not yet been sufficiently explored using neurophysiological or pharmacological approaches.^2,3,7,9,10^ Small trials of noradrenergic drugs in PSP led to overall heterogeneous clinical results and did neither include biomarkers nor cognitive or neuropsychiatric measures.^23–25^ To date, no clinical trials have specifically evaluated the efficacy of SNRI in PSP.

In this study, our first aim was the assessment of pupil-linked arousal deficits in PSP – ranging from alterations in baseline size and spontaneous pupil fluctuations to impaired stimulus-evoked responses – and their association with arousal and executive function, shedding light on the cognitive and behavioral consequences of ARAS dysfunction in PSP. Our second aim was to target ARAS dysfunction in PSP by administration of SNRI, modulating the serotonergic and noradrenergic neurotransmission within the ARAS. We hypothesize that SNRI application in PSP affects clinical symptoms and pupil-linked arousal, providing novel insights into the mechanistic and behavioral consequences of neuromodulatory deficits of the ARAS.

## Methods

### Participants

16 PSP patients (eight females, aged 70.81 ± 6.32 years) and 14 healthy age-matched controls (six females, aged 69.21 ± 4.81 years) were recruited at the *University Medical Center Hamburg-Eppendorf* (**Table 1**). There were no significant differences regarding age or gender distribution between groups. This study was conducted in agreement with the declaration of Helsinki (1967) and was approved by the local ethics committee (ethics approval number: 2020-10060-BO-ff). All participants gave their informed consent to participate.

**Table 1.** Demographic and clinical characteristics of PSP patients. Gender (M=male, F=female), age (years), probable PSP subtype (according to MDS-PSP criteria 2017)^1^, disease duration (years), PSP stage (range 1-5)^26^, Montreal Cognitive Assessment (range 0-30, MoCA), and PSP Rating Scale (range 0-100)^27^ scores are shown. PSP subtypes are defined as follows: PSP-RS – Richardson’s syndrome; PSP-P – PSP with predominant parkinsonism; PSP-PGF – PSP with progressive gait freezing; PSP-F – PSP with frontal predominance. When multiple subtypes are indicated (e.g., PSP-RS/-P), this denotes patients presenting overlapping features of the primary subtype and the secondary variant. Asterisks (*) indicate patients dropping out from the study.

| Case, Gender, Age range | Prob. Subtype | Disease duration | PSP stage | MoCA | PSP rating scale |
| --- | --- | --- | --- | --- | --- |
| 1, M, 76-80 | PSP-RS /-PGF | 2 | 3 | 27 | 20 |
| 2, M, 61-65 | PSP-RS | 4 | 4 | 28 | 39 |
| 3, F, 71-75 | PSP-P | 2 | 1 | 26 | 14 |
| 4, M, 66-70 | PSP-RS | 4 | 4 | 27 | 42 |
| 5, F, 61-65 | PSP-RS | 4 | 4 | 20 | 45 |
| 6, M, 61-65 | PSP-RS /-P | 3 | 2 | 16 | 44 |
| 7, M, 61-65 | PSP-PGF /-F | 4 | 3 | 22 | 33 |
| 8, F, 76-80* | PSP-F | 6 | 3 | 23 | 30 |
| 9, M, 76-80* | PSP-P /-F | 11 | 3 | 21 | 38 |
| 10, F, 66-70 | PSP-P | 4 | 4 | 26 | 37 |
| 11, F, 66-70 | PSP-P /-F | 12 | 4 | 14 | 38 |
| 12, F, 66-70 | PSP-RS /-P | 1 | 2 | 28 | 23 |
| 13, M, 71-75 | PSP-P /-PGF | 6 | 3 | 23 | 24 |
| 14, F, 66-70 | PSP-RS /-F | 3 | 3 | 24 | 28 |
| 15, F, 71-75 | PSP-P | 1 | 2 | 26 | 31 |
| 16, M, 81-85 | PSP-F | 8 | 3 | 11 | 47 |

All patients met the Movement Disorder Society (MDS) criteria for a probable PSP diagnosis. Seven patients fulfilled the criteria for Richardson’s syndrome (PSP-RS), while the remaining nine patients presented with variant PSP subtypes (vPSP), including PSP with parkinsonism (PSP-P), PSP with progressive gait freezing (PSP-PGF), and PSP with frontal predominance (PSP-F). Inclusion and exclusion criteria were adapted from the multicenter Prospective PSP trial (ProPSP). Additionally, patients were required to be able to repeatedly walk a distance of seven meters, using a walking aid if necessary.

### SNRI administration

All 16 included PSP patients initiated SNRI treatment with Venlafaxine after the baseline visit (**Figure 1a**), with gradual titration over 2–3 weeks. Treatment duration was ≥4 weeks to ensure adequate drug exposure. Assessments at both visits were conducted in the absence of dopaminergic medication. Two patients discontinued within 2 weeks due to adverse effects (dizziness and nausea in one; postural instability with increased falls in the other), leaving 14 patients for comparative analysis of clinical and cognitive outcomes. Follow-up assessments were performed after 5.6 ± 1.3 weeks at a mean SNRI dosage of 144.6 ± 28.9 mg.

**Figure 1.**
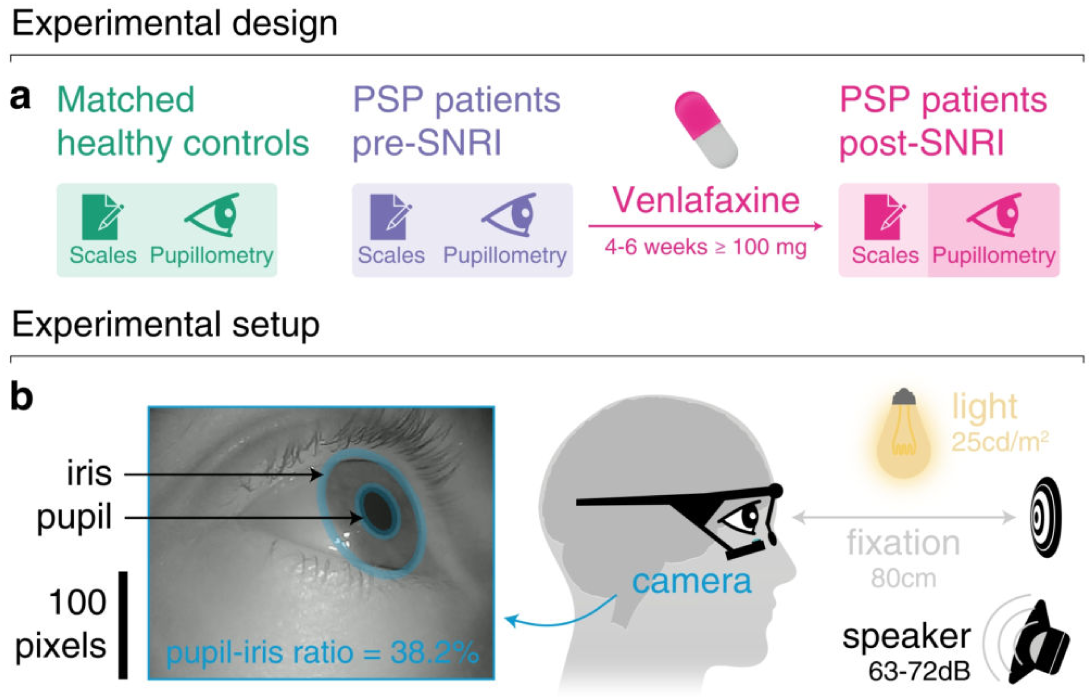
Study design. **a**, Design of the study: Pupillary dynamics and clinical scales in 14 PSP patients were assessed before and after chronic 4–6-week SNRI (serotonin-noradrenaline reuptake inhibitor) treatment and compared to 14 age-matched control participants. Note that recording quality was adequate for analyses in 11 (pupillometry at rest) and 9 (pupillometry during auditory sequence) participants. **b**, Experimental setup: Pupillometrics were measured by head-mounted goggles. To enable cross-session comparisons, pupil size was normalized relative to iris diameter.

### Clinical test and scores

For clinical characterization, rater-based assessments comprised motor evaluation by the PSP Rating Scale (PSPRS)^27^ and PSP clinical deficits scale (PSP-CDS).^28^ Cognitive evaluation was performed by phonemic verbal fluency (PVF), Trail making test (TMT) -A and -B and Montreal Cognitive Assessment (MoCA) at baseline and at follow-up visit. MoCA was adapted (*n* = 1), and TMT-B was conducted in oral (*n* = 3) instead of written form (WTMT-B), if the patient’s gaze was strongly impaired and confounded test performance. In that case, to enable comparability, the MoCA score was adapted by applying the factor ‘correct features / tested features’ (e.g., 19 out of 25 tested features correct = 0.76) to the standard total score of 30. The oral TMT B (OTMT-B) was converted by means of the WTMT-B/OTMT-B ratio, which was 2.3 for age 59-90 according to Mrazik et al.^29^

At both baseline and follow-up visits, participants completed the following self-report questionnaires, with assistance from a relative or research staff if needed: Geriatric depression scale (GDS), PSP quality of life scale (PSP-QoL),^30^ Starkstein Apathy scale (SAS) and Munich dysphagia test (MDT). For group comparison, healthy control subjects were assessed using MoCA, Trail Making Test A and B (TMT-A, TMT-B), phonemic verbal fluency (PVF), and GDS.

### Pupillometric recordings

For pupillometry measurements, participants were seated in a semi-dark room with constant luminance conditions (luminance 25.76 ± 0.7 cd/m^2^), in 80 cm distance from the wall, where a fixation cross on a white 21 cm x 29.7 cm rectangle was displayed (**Figure 1b**). Pupillometry data were captured by *Pupil Labs* (version 3.4.0) using eye tracking glasses (*Pupil Labs Core*, *Pupil Labs*, Berlin, Germany). The eye tracker was calibrated manually using a physical marker. Each eye was sampled with 125 frames per second. Data was recorded by the *LSL Lab recorder* (version 1.14.1b9). Pupillometry was performed under two different conditions.

#### Resting condition

This experimental block was designed to record spontaneous pupil fluctuations to capture baseline arousal dynamics. After calibration, pupil data were recorded in resting condition three times for 3.5 minutes each. The subjects were asked to fixate a fixation cross displayed on the wall.

#### Oddball condition

This experimental component was used to provide complementary task-evoked measures of cognitive processing and attention, allowing complementary insights particularly in cognitive function. Pupil data were recorded during an auditory oddball paradigm, in which a total of 360 stimuli was presented in approximately 15 minutes. The auditory, randomized sequence consisted of two different sounds (low-pitched and high-pitched sound) with two different stimulus probabilities, a rare stimulus with 25% and a frequent sound with 75% probability, in a within-group counterbalanced manner, but remaining the same intra-individually at the long-term follow-up. The sounds (duration 50 ms, volume adjusted to the subjects’ comfort (range: 63-72dB),^31^ jittered inter-stimulus intervals between 1.8-3.2 s) were created in *MATLAB R2021a*, each consisting of three frequencies (Low: [350, 700, 1400], High: [500, 1000, 2000], Hz) with waning and waxing phases (7 ms). Sounds were set up by *MATLAB*, *Psychtoolbox* (version 3.0.17), delivered through a small speaker (*JBL Go 2*) in front of the subject and recorded by the *Lab Streaming Layer (LSL) Lab* recorder. To heighten attention, the experiment leader announced at the start that a question would follow the sound sequence. Participants were asked to maintain fixation and to listen carefully to the sounds. The recording contained two small breaks (after 5 and 10 minutes), during which subjects could release fixation and relax. After the sound sequence, they were asked to estimate the probability of the rare sound.

### Preprocessing of pupil data

Two-dimensional pupil data in pixels provided by *Pupil Labs* was chosen for analysis. Preprocessing was performed with *MATLAB R2023b* by sorting original samples, resampling at a continuous sampling rate of 125Hz, blink, artifact and noise rejection, interpolation and filtering (low pass butter filter with a cut-off of 10 Hz, high pass butter filter with a cut-off of 0.01 Hz in the oddball condition). The pupil size was normalized by dividing the pupil diameter [pixels] by the iris diameter [pixels] resulting in pupil size in % of iris size. This approach minimizes variability from individual biological traits and camera geometry, providing a relative, dimensionless measure comparable across subjects and experimental setups.^32^ The analysis was focused on one “best” eye defined as the one with more valid segments (resting condition) or valid trials (oddball), remaining constant across baseline and follow-up sessions.

### Pupil size and spontaneous pupil fluctuations at rest

The average pupil size and its standard deviation were computed by averaging across the time course of each recording at rest and then averaged on subject level. Analysis of power-frequency spectra was performed in *MATLAB R2023b*. Data segments shorter than 24 s were discarded. Remaining data segments were detrended (the best straight-line fit linear trend was removed). Then, a Fast Fourier Transformation (FFT) was computed for each data segment using Welch’s method with a window of 20s and an overlap of 15 s. Power-frequency spectra were weight-averaged first across segments of one recording, then across recordings, resulting in one power-frequency spectrum per session.

The area under the curve (AUC) of the power-frequency spectra was calculated at subject level relative to a power curve fitted to the data at group level (AUC = integer of subject power spectral density - integer of fitted power curve) comprising the frequency range of interest from 1/16 to ½ Hz. Last, power-frequency spectra as well as area under the curve values were group averaged.

The power-frequency spectra were further analyzed by means of spectral parametrization in *Python* (specparam toolbox, https://specparam-tools.github.io/),^33^ decomposing the signal into an aperiodic component reflecting the 1/*f* background and a periodic component representing the pupillary hippus, a rhythmic oscillation of pupil size.^34,35^ This allowed us to quantify the aperiodic component as well as to extract the hippus peak frequency and power.

### Stimulus-evoked pupil dilation responses

The pupil size time series of each subject and session was epoched with a window from −0.4 to 3 s with respect to stimulus onset to capture the pupil dilation response. Pupil size was then baseline-corrected by subtracting the pupil baseline size averaged from −0.2 to 0 s. Epochs containing NaN values were discarded. Pupil size epochs of all trials, of frequent stimulus- and rare stimulus trials were averaged at subject level.

Peaks in pupil dilation response were located within a defined pupil size window from 0.5 to 2 s (*MATLAB* “max” function), resulting in values for peak amplitude and latency. Next, the area under the curve was calculated for the subjects’ pupil dilation response with a window from 0 to 1.8 s (*MATLAB* “trapz” function).

The first derivative of pupil size was calculated (*MATLAB* “diff” function), resulting in a measure indicating the velocity of pupil size change [% of iris size per second]. Peaks in the derivative of pupil size were located to detect the maximum speed in pupil dilation and its latency.

A logistic regression model was fitted on the pupil dilation responses, with the pupil time series as predictor and the stimulus identity as a dependent variable for each oddball trial, resulting in relative log-likelihood as a measure for model performance in predicting stimulus identity based on changes in pupil size.

The surprise pupil response was calculated at subject level by subtracting the mean pupil response to frequent stimuli from the mean pupil response to rare stimuli. Here, the first derivative of the signal was calculated after applying an additional low-pass filter with a cut-off of 3 Hz.

### Eye blinks and related changes in pupil size

Since blink frequency is known to be decreased in PSP patients^36^ and a so-called blink-locked pupil response — a constriction and consecutive dilation following eye blinks — has been described,^37^ the following analyses regarding eye blinks and their effect on pupil size were performed to rule out a bias resulting from differences in eye blinking.

Artifacts which were detected in the preprocessing and fulfilled criteria of eye blinks (duration 160-600 ms) were transformed into a binary vector indicating the occurrence of blinks. For an overall blink rate [blinks per minute], the total number of blinks was divided by recording duration [min]. To investigate the relationship of blinking and stimulus presentation, the blink vector was epoched from −3 to 3 s with respect to stimulus onset and a peri-stimulus time histogram was computed resulting in a blink rate [blinks per second]. To evaluate if the blink rate was increased after stimulus presentation, the average blink rate from 0 to 1 s was compared to the blink rate baseline averaged from −1 to 0 s with respect to stimulus onset.

### Modulation of pupil response by theoretical surprise

According to seminal contributions in the field, the brain constantly attempts to infer the statistical structure of its environment.^38^ Correspondingly, computational models of the inference process developed to account for this process capture brain activity in a variety of contexts.^39^ To quantify the extent to which pupil responses reflect theoretical surprise levels, we used a previously proposed model resting on Bayesian inference,^40^ which was shown to explain brain activity in similar experimental situations (binary auditory sequences, passive listening).^41^ Specifically, the model estimates the matrix of transition probability between sequence events and compares newly received observations against existing beliefs to yield a theoretical surprise response. The model is parameterized with an integration timescale which controls the exponential forgetting decay with which past observations are progressively discarded from the inference. Local integration timescales lead to fast forgetting and large fluctuations in surprise levels while global integration timescales lead to slow forgetting and small fluctuations in surprise levels due to progressive convergence of inferred probabilities towards true, generative probabilities. Surprise levels from the model — as estimated using different integration timescales (from 1 to 100, logarithmically spaced) — were used to predict pupil responses to auditory stimuli at different latencies with a linear regression. The resulting proportion of variance explained was compared across experimental groups.

### Statistical analysis

Statistical analysis of clinical scores, gait variables, pupil and blink metrics was performed in *MATLAB R2023b*. Shapiro-Wilks test was performed to check the data for normal distribution, resulting in partly not normally distributed data. Therefore, nonparametric statistical tests were chosen, Mann-Whitney U tests for inter-group-comparison (PSP vs HC), and Wilcoxon signed rank tests for within-group-comparison of PSP at baseline (BL) and PSP at follow-up (FU).

Statistical analysis of continuous data was performed in *MATLAB R2023b* using *Fieldtrip*’s toolbox for cluster-based permutation tests,^42^ which is widely used for statistical analysis of one- or multidimensional data as it addresses the multiple comparisons problem. The pupil dilation response and its derivative were compared between groups by means of cluster permutation testing from 0 to 1.8 s with respect to stimulus onset with alpha = 0.025 and the number of permutations set to 10 000 for independent testing of HC and PSP, while all possible permutations were performed when testing dependently between PSP sessions.

To establish for each group separately if a significant surprise response was present, a cluster permutation test of the signal against zero was performed with alpha = 0.025 and all possible permutations. To test the logistic regression model performance, the log-likelihood was tested by means of cluster permutation test against the baseline mean (from −0.2 to 0 s) for each group.

To compare the surprise pupil response and its derivative between groups, cluster permutations tests were performed as described above for the pupil dilation response.

## Results

The aim of the experimental series was to investigate pupil-linked arousal dynamics in progressive supranuclear palsy (PSP). First, we characterized pupillary dynamics compared to healthy controls and examined their association with clinical parameters as gait (see supplementary material) and cognitive function to assess their value as indirect markers of clinical function. Second, we evaluated the effect of the neuromodulatory SNRI treatment on pupillary dynamics and related clinical outcomes, thereby underscoring the close link between pupil behavior and clinical manifestations.

### SNRI treatment improves executive function and quality of life

Overall, PSP patients performed worse than healthy controls across all clinical measures, with only circumscribed improvements observed under SNRI treatment (**Figure 2**).

**Figure 2.**
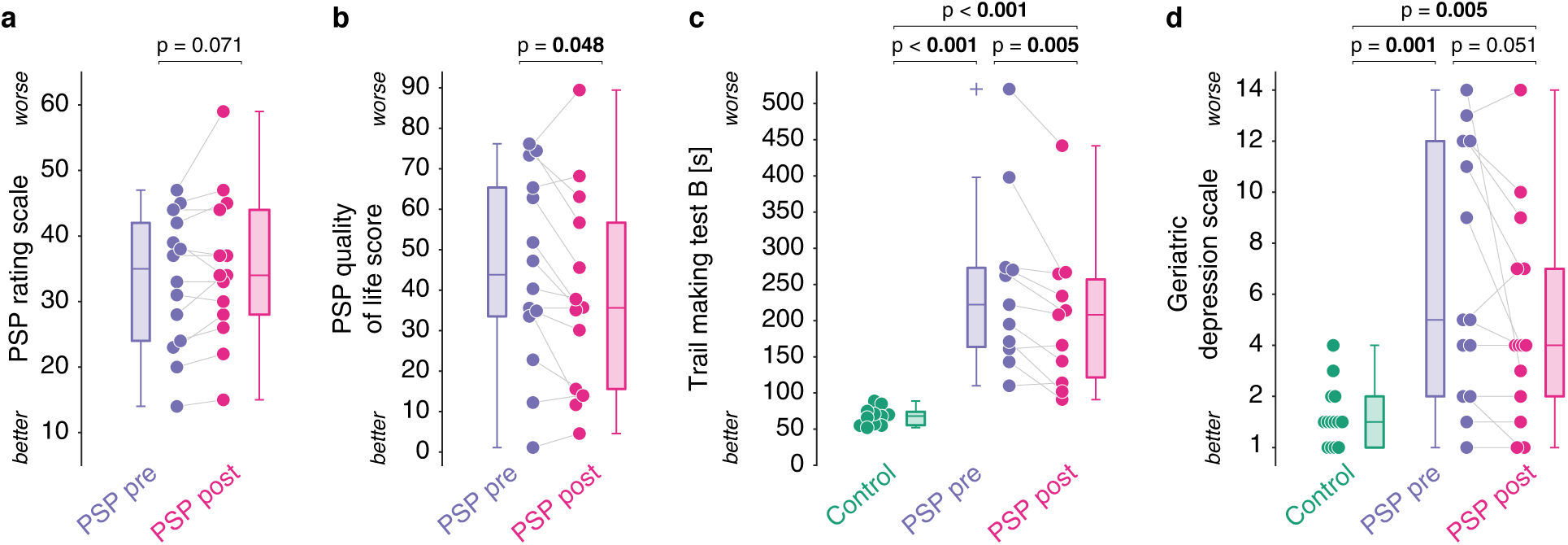
Clinical scores and questionnaires. Clinical and neuropsychological measures in PSP before and after SNRI treatment. Boxplots (median, whiskers) with scattered dots representing individual mean values. Colors indicate PSP at baseline (purple), at follow-up at 4-6 weeks (pink), and healthy controls (green, where applicable). (a) PSP Rating Scale. (b) PSP Quality of Life Score. (c) Trail Making Test B (TMT-B). (d) Geriatric Depression Scale.

The *PSP rating scale* did not significantly change between baseline and the follow-up visit with SNRI therapy (*n* = 14, *p* = 0.071). Suitably, objective gait analyses did not reveal any motor improvement by SNRI therapy (see supplementary material). However, the *PSP-Quality of Life* score was significantly decreased at follow-up (38.78 ± 24.11) compared to baseline (45.10 ± 23.63), revealing an improvement of quality of life by treatment (*n* = 14, *p* = 0.048).

On the *Geriatric Depression Scale*, PSP patients scored significantly higher than healthy controls (HC) at both baseline (*n* = 14, *p* = 0.001) and follow-up (*n* = 14, *p* = 0.005), indicating more depressive features. Between patient visits, scores showed a slight, but nonsignificant decrease at follow-up with respect to baseline (BL: 6.71 ± 4.92; FU: 4.93 ± 4.05; *n* = 14, *p* = 0.051). Also, patient questionnaires evaluating dysphagia, apathy and activities of daily living were not significantly improved by SNRI therapy (MDS *n* = 14, *p* = 0.626; SAS *n* = 13, *p* = 0.565; SEADL *n* = 14, *p* = 0.25).

Regarding cognitive tests, PSP patients performed poorly at both visits when compared to HC. MoCA total scores as a general measure of cognitive performance were significantly lower in PSP at baseline (22.74 ± 5.48, *n* = 14, *p* < 0.001) and follow-up (23.3 ± 5.20, *n* = 14, *p* = 0.001) compared to HC, with no significant effect of SNRI therapy in PSP subjects at follow-up visit (*n* = 14, *p* = 0.511). In the phonemic verbal fluency (PVF) test, patients generated significantly fewer words than HC at both baseline and follow-up (PSP BL vs. HC, *n* = 14, *p* < 0.001; PSP FU vs. HC, *n* = 14, *p* < 0.001), whereas performance did not differ between PSP visits (*n* = 14, *p* = 0.779).

The TMT-A, mainly reflecting basic attentional and speed-related processes, was significantly different between HC and PSP at both visits (PSP BL vs. HC, *n* = 10, *p* = 0.00; PSP FU vs. HC, *n* = 10, *p* = 0.00), but again was not improved by SNRI therapy in PSP (PSP FU vs. BL, *n* = 10, *p* = 0.576). In TMT-B, which assesses sequential set-shifting as part of executive functions, PSP patients also performed significantly worse at both visits when compared to HC (PSP BL vs HC, *n* = 11, *p* < 0.001; PSP FU vs. HC, *n* = 11, *p* < 0.001). However, comparison of TMT-B outcomes of PSP at baseline (247.76 ± 120.31) and follow-up (204.17 ± 100.69) with SNRI revealed significant improvements, as they took less time to complete the test at follow-up visit (*n* = 11, *p* = 0.005). When calculating the TMT delta (TMT-B – TMT-A), which reduces the influence of processing speed and more selectively reflects executive control, significant differences between PSP and HC (PSP BL vs. HC, *n* = 9, *p* < 0.001; PSP FU vs. HC, *n* = 9, *p* < 0.001), as well as between PSP visits (*n* = 9, *p* = 0.012) persisted.

Thus, in summary, SNRI treatment improved selective clinical symptoms with particular effect on executive function and quality of life.

### Patients present smaller pupil size and abnormal pupil fluctuations at rest

Pupillary dynamics at rest in PSP were altered at baseline compared to healthy controls and partially normalized at follow-up (**Figure 3**). At baseline, PSP patients exhibited smaller average pupil size and lower variability compared to healthy controls (HC) (size: *n* = 11, *p* = 0.088; SD: *n* = 11, *p* = 0.01). At follow-up, both measures increased significantly compared to baseline (size: *n* = 11, *p* = 0.003; SD: *n* = 11, *p* = 0.01) and were no longer significantly different from HC (size: *n* = 11, *p* = 1.00; SD: *n* = 11, *p* = 0.131). Resting-state power spectral density (PSD) of pupil fluctuations was reduced across the frequency spectrum in PSP. Cluster permutation testing revealed two significant clusters at baseline compared to HC — at very low frequencies up to 1/8 Hz (*n* = 11, *p* = 0.014) and around 1/4–1/2 Hz (*n* = 11, *p* = 0.002). These clusters remained significant at follow-up (*n* = 11, *p* = 0.022; *n* = 11, *p* = 0.016), while overall PSD increased slightly without reaching significance between PSP sessions. Spectral parametrization confirmed a lower offset of the aperiodic component in PSP at both visits compared to HC (BL: *n* = 11, *p* = 0.001; FU: *n* = 11, *p* = 0.007), whereas the aperiodic exponent did not differ (*n* = 11, *p* > 0.26).

**Figure 3.**
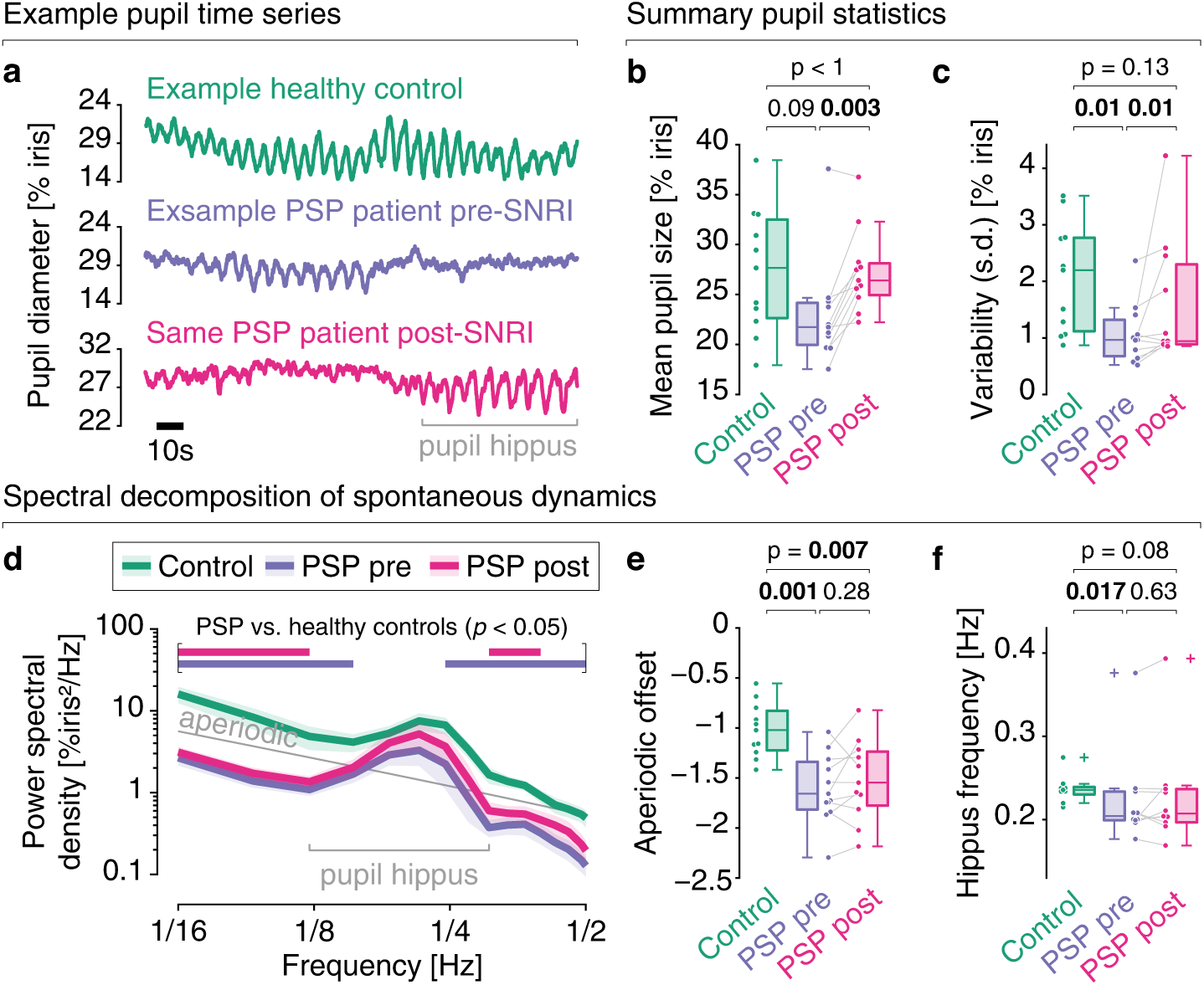
Impairments of spontaneous pupil dynamics in PSP and their partial recovery with SNRI. **a,** Example pupil time series at rest for healthy controls (green), PSP at baseline (purple) and at follow-up (pink). **b,** Mean pupil size averaged across pupil time series. **c,** Standard deviation of pupil time series as a measure of pupil variability. **d,** Power spectrum across frequencies of healthy controls (green), PSP at baseline (purple) and at follow-up (pink). The solid line represents the mean, while the shaded area equals the standard error of the mean. The thin grey line is a curve fitted to the power-frequency spectra from all groups and used for AUC computation. Bars at the bottom indicate statistically significant clusters (*p* < 0.05) from cluster permutation testing. **e,** Offset of aperiodic component extracted by spectral parametrization of power-frequency spectra. **f,** Hippus frequency from periodic components extracted by spectral parametrization of power-frequency spectra.

Hippus oscillations were present in 10/11 PSP patients and 9/11 HC. Hippus frequency was significantly lower in PSP at baseline (*n* = 11, *p* = 0.017) but normalized at follow-up (*n* = 11, *p =* 0.079) with no significant difference between PSP sessions (*n* = 11, *p* = 0.625). The area under the curve (AUC) around the average hippus frequency was reduced in PSP at both visits compared to HC (BL: *n* = 11, *p* = 0.001; FU: *n* = 11, *p* = 0.006) with no significant change between sessions (*n* = 11, *p* = 0.577). These AUC differences were largely driven by the aperiodic component, as spectral parametrization revealed no significant changes in the periodic (hippus) power.

In summary, PSP is characterized by reduced pupil size, variability, and PSD at baseline, with partial recovery at follow-up, with alterations in the aperiodic component mainly driving differences in pupillary oscillatory dynamics.

### Stimulus-evoked pupil dilation responses (PDR) were reduced in PSP and partially recovered by SNRI

Given that pupil size is closely linked to cognitive processing, we aimed to assess whether cognitive modulation of pupil dynamics is altered in PSP and whether it can be influenced by SNRI treatment. First, we focused on the well-described pupil dilation response (PDR) to sensory binary auditory sequences with biased uneven tone probabilities (**Figure 4a**).^41,43,44^ We found no group differences regarding the estimated probability for the rare stimulus reported by PSP patients and HC at the end of the auditory sequence (**Figure 4b**). PSP patients presented smaller evoked PDR than HC, which appeared slightly increased at follow-up compared to baseline (**Figure 4d**). However, cluster permutation tests from 0 to 1.8 s as well as the area under the curve for the same time window did not reveal significant differences between groups or sessions.

**Figure 4.**
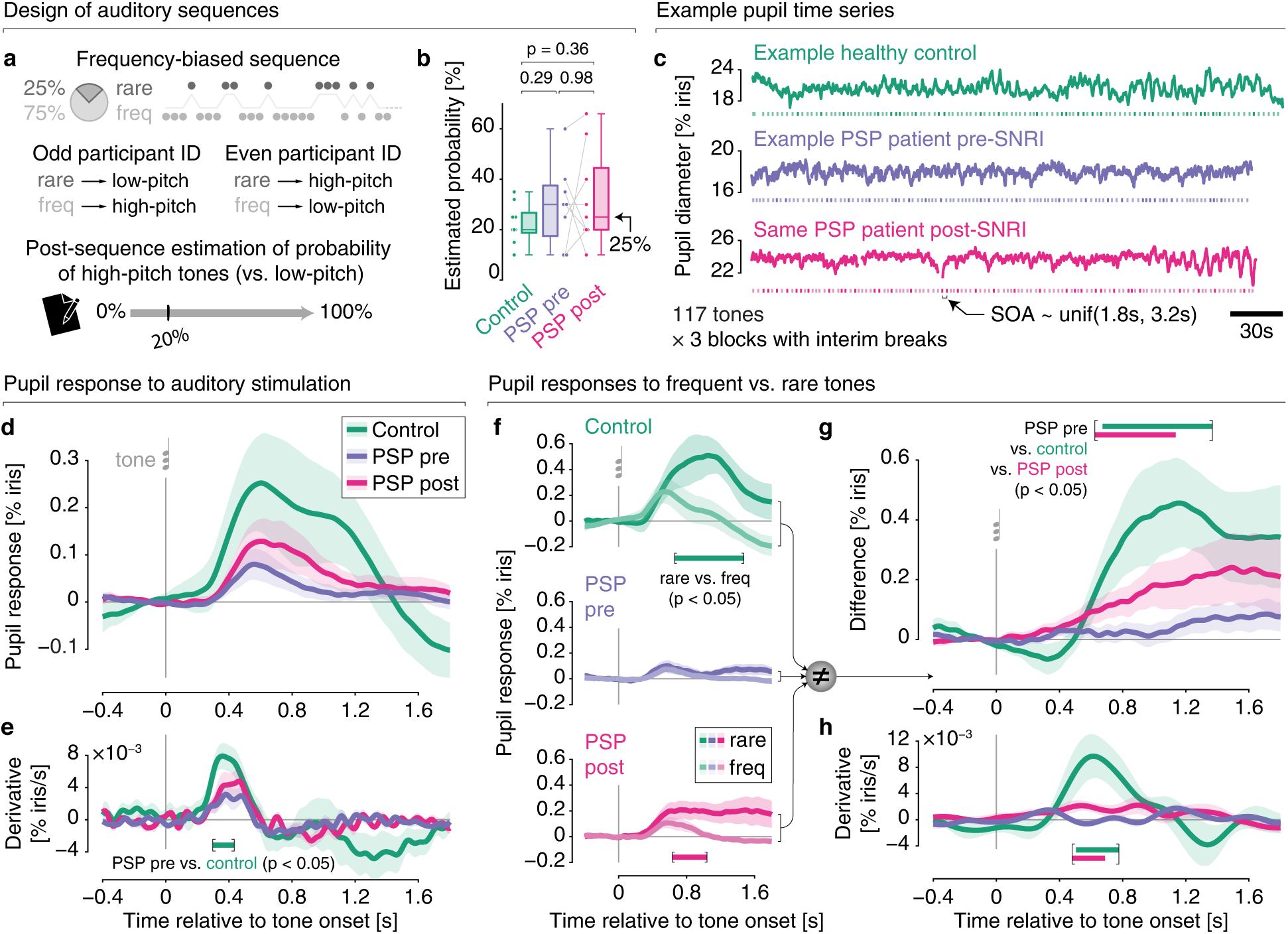
Impairments of pupil responses to surprising auditory events in PSP and their partial recovery with SNRI. **a,** Experimental design of an auditory sequence for the oddball paradigm, consisting of a high- and a low-pitched tone, one of which is assigned as a rare stimulus with a probability of 25% in a counterbalanced manner. **b,** Boxplot of the estimated frequency with which the rare stimulus occurred, estimated by the participant after the end of the sequence. **c,** Example pupil time series and sound sequence (below the pupil trace) from the oddball paradigm for each group: healthy controls (green), PSP at baseline (purple) and at follow-up (pink). **d,** Overall pupil dilation response: Mean (colored line) and standard error of the mean (s.e.m.; shaded area) for each group. **e,** First-order derivative of pupil dilation response: Mean (colored line) and s.e.m. (shaded area) for each group. The bar indicates a significant cluster (*p* < 0.05) from cluster permutation testing. **f,** Pupil dilation response toward rare vs. frequent stimuli for each group: Mean (colored line) and s.e.m. (shaded area) for PDR to both rare (darker color) and frequent tone (brighter color). Bars at the bottom indicate significant clusters (*p* < 0.05) from cluster permutation testing. **g,** Surprise pupil response (PDR rare - PDR frequent): Mean (colored line) and s.e.m. (shaded area) for each group. Bars at the top indicate significant clusters (*p* < 0.05) from cluster permutation testing. **h,** First derivative of pupil surprise response: Mean (colored line) and s.e.m. (shaded area) for each group. Bars in the bottom indicate significant clusters (*p* < 0.05) from cluster permutation testing.

Comparing the peaks in PDR showed significantly smaller peaks in PSP at baseline than in HC (HC *n* = 9, PSP *n* = 8; PSP at BL vs. HC, *p* = 0.011; PSP at FU vs. HC, *p* = 0.094; PSP FU vs. BL, *p* = 0.055). going along with a significantly decreased peak latency in PSP at baseline compared to HC (HC *n* = 9, PSP *n* = 8; PSP at BL vs. HC, *p* = 0.046; PSP at FU vs. HC, *p* = 0.114; PSP FU vs. BL, *p* = 1).

Since the speed of pupil size change was reported to correlate more specifically to LC activity,^16,22^ we analyzed the first-order derivative of the PDR (**Figure 4e**). In line with the difference in steepness of the pupil response curves, analyses confirmed a significantly smaller pupil derivative in PSP at baseline than in HC (*n =* 9, *p* = 0.045), while there were no significant clusters detected when testing for differences between PSP at follow-up and HC or between PSP sessions.

### Surprise pupil responses are dampened in PSP and partially recovered by SNRI

In addition to inspecting the overall PDR to sensory stimuli, we moved to the study of the surprise-modulated pupil response, putatively indexing higher-level cognitive processing. In line with past findings,^19,43^ the PDR to rare sounds was larger than to frequent sounds in PSP patients and HC (HC: *n =* 9, *p* = 0.01; **Figure 4f**). However, in PSP a significantly greater pupil response to rare sounds was only evidenced at follow-up (PSP at FU: *n* = 9, *p* = 0.031; **Figure 4f**), while there was no cluster indicating a surprise pupil response at baseline. Group comparison of the surprise pupil response (**Figure 4g**) showed a significantly smaller response in PSP at baseline compared to HC (*n* = 9, *p* = 0.013) as well as an increased surprise response in PSP at follow-up compared to baseline (*n* = 9, *p* = 0.021). PSP at follow-up and HC did not differ significantly. To assess not only alterations in amplitude and latency, but to investigate how far the pupil responses in PSP are modulated by stimulus identity, a model was fitted to the pupil responses. Group comparison showed a weaker model performance when attempting to decode stimulus identity from the pupil response in PSP patients, especially at baseline (**Supplementary figure 1**). We further analyzed the derivative of the surprise response (**Figure 4h**) and found a significant cluster indicating a smaller derivative in PSP at baseline compared to HC (*n* = 9, *p* = 0.012), as well as a cluster indicating a significantly increased derivative in PSP at follow-up compared to baseline (*n =* 9, *p* = 0.023). There were no significant clusters when comparing PSP at follow-up and HC.

Importantly, the pupil does not respond to rare and frequent stimuli in a static manner. Instead, the amplitude of the PDR is a complex function of the immediate history of past stimuli. For instance, the longer the history of frequent stimuli preceding the rare stimulus, the more the PDR to rare stimuli tends to be amplified. These complex sequence effects are normative under the assumption that the brain continuously infers local statistical dependencies in the environment, and are consequently well accounted for by probabilistic inference models.^41^ To determine whether the brain’s probabilistic inference computations — as indexed by subtle trial-to-trials fluctuations in the PDR — were disrupted in PSP and possibly partially rescued by SNRI administration, we relied on a previously proposed model of sequence processing that predicts subtle variations in theoretical surprise levels (**Figure 5a,b**),^40^ and used it to predict the time-resolved pupil fluctuations during auditory sequence presentation (**Figure 5c,d**). Analysis of the model’s quality-of-fit (i.e., proportion of explained variance) revealed that pupillary reflections of probabilistic computations were severely impaired in PSP patients, by comparison to HC (*n* = 9, *p* = 0.024; **Figure 5e**). This however was improved after treatment where no significant differences could be detected in comparison to HC (*n* = 9, *p* = 0.14).

**Figure 5.**
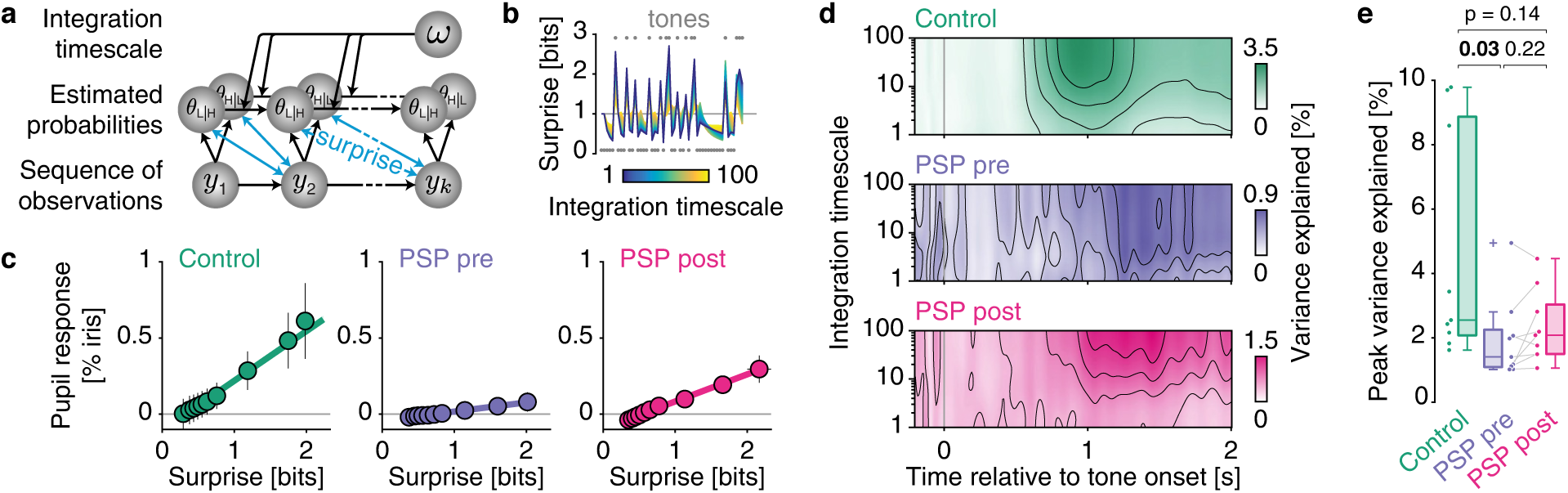
Impairment of pupil-reflected probabilistic inference in PSP and its partial recovery with SNRI. **a,** Computational model of sequence processing. Transition probabilities are inferred from a sequence of observations and theoretical surprise is obtained by comparing inferred probabilities against received observations. An integration timescale parameter controls the amount of exponential forgetting in the model. **b,** Dynamics of theoretical surprise according to different integration timescale in the model. **c,** Linear scaling of the pupil response according to theoretical surprise in each group (10 equally sized bins). Surprise is obtained from the model with an integration timescale yielding the strongest proportion of variance explained (on an individual basis). Pupil response is taken at a latency yielding the strongest proportion of variance explained (on an individual basis). **d,** Proportion of variance explained as a function of both pupil response latency and integration timescale for each group. **e,** Peak proportion of variance explained (across pupil response latency and integration timescale) in each group (*p*-values correspond to a rank-sum test).

### Eye blink frequency did not explain pupil differences

We ended by considering the possibility that PDR differences between groups and conditions could be confounded by a difference in eye blink frequency or occurrence of stimulus-related eye blinks. Blink rate was significantly reduced in PSP (PSP BL vs. HC, *n* = 9, *p* = 0.019; PSP FU vs. HC, *n* = 9, *p* = 0.011), without differences between sessions (FU vs. BL, *n* = 9, *p* = 1.000). An increased blink rate after stimulus onset was observed in HC (*n* = 9, *p* = 0.039), but not in PSP patients (*n =* 9; PSP BL, *p* = 0.57; PSP FU, *p* = 0.73). For all groups and conditions there were no significant differences in PDR before and after excluding trials containing blinks (**Supplementary figure 2**). We therefore conclude, the observed PDR differences are not driven by blink-associated pupil dynamics.

## Discussion

In this study, PSP patients presented with characteristic clinical features including cognitive-executive, motor, and depressive symptoms, which were accompanied by specific changes in pupillometry. PSP patients exhibited smaller pupil size, diminished pupil responses to auditory stimuli and impaired surprise pupil responses at baseline compared to controls, which may be attributed to a deficient ARAS in PSP. Chronic SNRI administration over 4-6 weeks led to a significant, yet selective improvement of executive functions which was associated with increased pupil size at rest, enhanced pupil size dynamics in both resting and oddball condition, with emphasis on a partial recovery of surprise pupil responses, which we propose to reflect re-established ARAS activity. To our knowledge, this is the first study to modulate ARAS activity in PSP using an SNRI and to non-invasively monitor the resulting clinical and neurophysiological effects via pupillometry. As a pilot study in PSP, our work was limited by the small sample size, open-label design with potential placebo effects and short duration of observation of 4-6 weeks.

The ARAS is a distributed network of brainstem nuclei and projections that sustain cortical arousal, modulate attention and autonomic functions. It comprises noradrenergic neurons of the locus coeruleus (LC), serotonergic raphe nuclei, cholinergic pedunculopontine (PPN), laterodorsal tegmental nuclei, dopaminergic neurons of the ventral tegmental area, and parabrachial glutamatergic inputs.^5^ These structures send projections to thalamic relay and intralaminar nuclei, the hypothalamus, basal forebrain, and cortex, forming both thalamic and extrathalamic routes that maintain wakefulness and responsiveness.^45^ Of particular interest is the LC in the dorsolateral pontine tegmentum, which provides widespread noradrenergic projections to cortex, hippocampus, thalamus, basal ganglia, cerebellum and spinal cord.^46–48^ Its tonic and phasic firing patterns support sustained attention and vigilance.^49^

In PSP, ARAS structures are among the earliest and most severely affected regions.^50,51^ Tau pathology in PSP seems to be initiated in brainstem nuclei, leading to their degeneration and contributing to hallmark symptoms such as postural instability, oculomotor dysfunction, and cognitive impairments.^4,51^ Neuroimaging studies have demonstrated disrupted white matter tracts connecting ARAS regions to cortical areas, correlating with the severity of attentional and executive dysfunctions observed in PSP patients.^10,52^ Additionally, positron emission tomography (PET) scans revealed reduced metabolic activity in ARAS-related regions, further implicating this system in PSP pathophysiology.^51^

So far, there is no conclusive evidence that ARAS degeneration differs between PSP subtypes.^1^ Data suggest that the ARAS is affected across all PSP variants, although the degree and timing of degeneration may vary and contribute to clinical heterogeneity.^3,53,54^ We found impaired pupil dynamics and the response to SNRI to be variable across patients. While a larger sample is needed to further investigate the observed pupil deficits, the PSP spectrum could also be leveraged to explore potential correlations between the distribution of tau pathology and the degree of pupil impairment. Further studies are needed to better assess the clinical benefit of noradrenergic treatment and, ideally, to identify predictive markers of responsiveness to SNRI.

The strong link between ARAS and pupil dynamics has been revealed by neuroimaging studies in health^14,19^ and disease.^44,55^ In PSP, pathologically reduced baseline pupil size, smaller pupil constriction and dilation responses in PSP patients during a video-based eye-tracking task were considered as a read-out of impaired ARAS activity.^55^ Moreover, other features of pupillometry such as the reduced blink rate and the absence of stimulation-induced blinks in PSP were interpreted as changed brainstem activity.^56,57^ In our experiment, we also interpret the pathologically impaired pupillary responses in PSP patients at baseline as an indirect indicator of ARAS deficiency prior to SNRI treatment.

We assessed the effect of SNRI treatment on clinical measures and pupillometry in PSP. After chronic SNRI administration of 4-6 weeks, we observed a circumscribed clinical improvement in cognitive executive function measured by the TMT-B and an increased quality of life, whereas there was no effect of SNRI therapy on gait and other PSP motor symptoms and only partial, but non-significant effects on depression and apathy. SNRI therapy induces a dose-dependent modulation of neurotransmitter systems that may differentially affect motor, limbic and cognitive domains. In the context of PSP, the LC, with its role in arousal and attention, suggests that SNRI’s action on noradrenaline pathways could be particularly beneficial for executive functions, which are often impaired in PSP patients. This hypothesis is supported by findings that SNRI treatment can reduce deficits in executive control of attention in patients with major depressive disorder.^58^

We propose that the lack of SNRI effect on gait and motor symptoms in PSP might be due to the differential vulnerability and functional roles of ARAS efferents and other motor-related circuits. Primary motor symptoms and gait disorder in PSP may not be driven by noradrenergic deficits alone, but result from degeneration of basal ganglia nuclei and midbrain structures critical for vertical gaze and postural control.^51^ These regions are only partially innervated by noradrenergic LC projections.^18,45^ Therefore, boosting LC signaling via SNRI therapy primarily affects cortical arousal and executive networks, not the downstream basal ganglia motor circuits controlling gait and posture, which also involve cholinergic, dopaminergic, and glutamatergic pathways.^4^ The SNRI’s pharmacological action on norepinephrine and serotonin may not restore these damaged circuits. Thus, SNRI therapy selectively enhances residual noradrenergic signaling in cortical and attentional networks,^6^ improving cognition in PSP. Its lack of effect on gait and motor symptoms may reflect a functional dissociation and independence of motor circuits from the noradrenergic LC–ARAS efferents.

Astonishingly, we failed to observe significant effects of SNRI therapy on apathy and only partial, but non-significant effects on depression in PSP patients. One explanation might be the short duration of SNRI wash-in of 4-6 weeks, since effects might increase even at 8 weeks.^59^ Another hypothesis would be a differential modulation of the LC and raphe nuclei by the SNRI, with more accentuated modulation of the LC. The raphe nuclei, primarily serotonergic, are also affected by tau pathology in PSP, though to a lesser extent than the LC.^2^ The differential involvement of the LC and raphe nuclei in PSP suggests that the noradrenergic system primarily mediates the cognitive symptoms, while the serotonergic system is more closely associated with mood and behavioral disturbances. This distinction might be supported by studies showing varying patterns of degeneration in these brainstem nuclei in PSP patients.^2^ Understanding the differential roles of these systems is crucial for developing targeted therapeutic strategies for PSP for distinct aspects of the disease.

This is the first study to assess the effect of SNRI on pupillary dynamics in PSP. In healthy volunteers, single therapeutic doses of SNRI increased resting pupil diameter, prolonged latency, reduced amplitude, and shortened recovery of the pupillary light reflex, reflecting central parasympathetic inhibition and sympathetic potentiation via noradrenaline reuptake blockade, with concentration-dependent noradrenergic effects of the SNRI, including rapid tolerance of pupillary light reflex parameters.^60–62^ We analyzed the pupillary phasic “surprise response” elicited by rare surprising stimuli, which is strongly linked to transient activation of the LC–noradrenaline system.^16–18,63^ Phasic bursts of LC activity occur in response to novel, salient, or surprising events, and these bursts are mirrored by rapid pupil dilations.^16,17^ In healthy, elderly subjects, the pupil response to auditory stimuli, and its ability to differentiate between rare and frequent tones, was reduced compared with younger and middle-aged individuals,^64^ in parallel with age-related changes in LC.^65^ Here, we observed decreased sound-evoked pupil responses in PSP compared with those of age-matched, elderly, healthy subjects. These group differences were particularly pronounced when considering not only the basic response to auditory stimuli, but also the contrast between responses to rare versus frequent tones. Here, PSP patients exhibited an impaired surprise pupil response at baseline; specifically, the rare sound did not elicit a significantly larger response than the frequent sound. Because the LC is particularly vulnerable in neurodegenerative diseases such as PSP, the magnitude and timing of the surprise-evoked pupillary response may serve as a particularly useful functional biomarker for LC integrity and noradrenergic signaling.^66^ Importantly, this biomarker is specific to phasic noradrenergic modulation rather than serotonergic or cholinergic systems, which contribute more tonically to baseline pupil size and slow fluctuations.^16^ We observed that SNRI therapy preferentially enhanced the pupillary surprise response in PSP, reflecting phasic LC signaling. Thus, the prefrontal cortical noradrenergic efferents of the ARAS may be preferentially stimulated, leading to improved executive function and attentional performance.

For future investigation specific neuroimaging techniques such as neuromelanin-sensitive MRI sequences to visualize LC cell bodies and PET tracers to assess noradrenaline transporter density display a valuable tool to gain insight into the alterations of the noradrenergic LC system in PSP. The relations between imaging findings and clinical data as well as pupillometry data should further be explored, complementing the described pupil deficits. Non-invasive measurement of pupil size displays a very promising biomarker for patients with neurodegenerative diseases, as we could evidence alterations in pupil-linked arousal in PSP patients – in line with those recently described in Alzheimer’s disease patients^44^ – and their partial responsiveness to SNRI therapy.

## Conclusion

For the first time, we describe alterations in pupil-linked arousal in PSP patients, manifesting as decreased baseline pupil size, slightly reduced pupil responses and markedly diminished surprise pupil responses, accompanied by cognitive impairment and depression. SNRI treatment partially rescued the impaired pupil metrics in PSP, along with an improvement in executive functions and quality of life. In summary, we report a close relationship between the observed pupillometry metrics and the symptoms in PSP as well as their partial responsiveness to SNRI treatment, paving the way for the use of pupillometric measures as a potential biomarker.

## Data availability

The datasets generated for this study are available on request to the corresponding author.

## Data Availability

The datasets generated for this study are available upon reasonable request to the corresponding author.

## Acknowledgment

We would like to thank the patients and their caregivers, as well as the healthy control persons, for their participation in this project.

This study was funded by the DFG (German Research Foundation), SFB 936, project C8 (M. Pötter-Nerger, C. Moll) and A7 (T. Donner).

M.M. is supported by a postdoctoral fellowship from the *Humboldt Stiftung* and by the *Fondation Bettencourt Schueller*.

