## Supplementary Material for "SNRI-driven recovery of impaired pupil dynamics in Progressive Supranuclear Palsy"

#### Supplementary figures

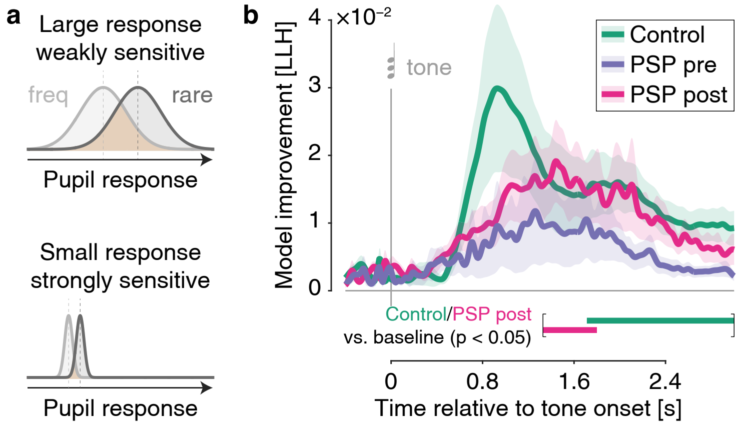


**Supplementary figure 1 | Reduction in the pupil-encoding of stimulus identity in PSP and its partial recovery by SNRI**

**a**, Example scenarios for SNRI-driven improvement of pupil response to rare auditory events in PSP: The amplitude of the pupil response is enhanced but remains weakly modulated by stimulus identity (top); or the amplitude of the pupil response remains small but becomes more strongly modulated by stimulus identity (bottom). Thus, two pupil phenomena can co-occur: A change in amplitude (see Figure 4) and a change in modulation by stimuli (see panel b).

**b**, Time-resolved performance of a logistic regression model predicting stimulus identity from the pupil response at different latencies. Model performance is quantified as log likelihood (LLH) relative to a null model without pupil response as predictor. Bars at the bottom indicate significant clusters ($p < 0.05$) from cluster permutation testing against baseline.

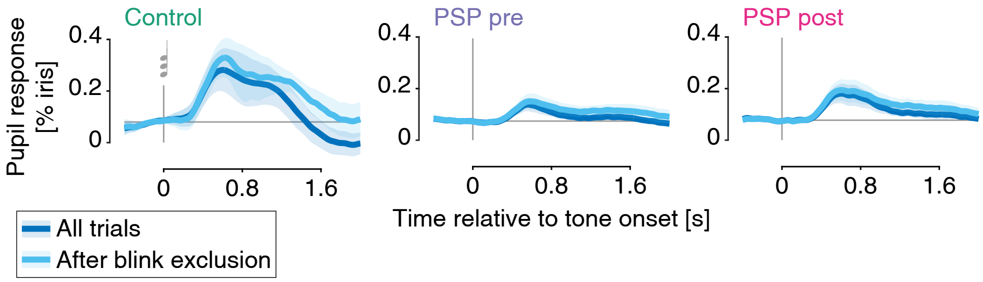


**Supplementary figure 2 | Pupil response after eye blink rejection**

Pupil response to tones (as in Figure 4d) across groups and conditions before and after rejecting trials entailing an eye blink.

### Supplementary material

*Gait analysis.* Two-dimensional spatiotemporal gait parameters were obtained by means of a pressure-sensitive carpet of 6.9 m length and *GAITRite* software version 4.8.6c7. If needed, patients could use a walking aid throughout the gait analysis. For safety reasons, a lab member was placed next to unsteady patients to prevent falls. Gait analysis was performed in three different conditions: at preferred speed, at maximal speed and while serially subtracting 7 (dual task). For each condition at least three walks were recorded. Using *MATLAB 2023b*, spatiotemporal gait variables were extracted and averaged per subject for each condition. As a measure of gait asymmetry, the asymmetry index (AI) was calculated for step length (AI = ratio of longer step length/shorter step length). Gait variability was assessed by calculating the coefficient of variability (CV) of step length and step time (CV = standard deviation / mean value × 100).

*Impairment of gait in PSP patients.* Gait impairment is a central symptom of PSP and is associated with cognitive dysfunction such as deficient attention or executive dysfunction.[67–70] For instance, an increased cognitive load – such as performing a task while walking – can worsen gait performance, an effect that has been found to be particularly pronounced in PSP.[67,69]

We analyzed data from objective gait analysis, comparing the PSP patients' gait performance with the gait of HC in three different conditions: walking 1. at preferred speed, 2. at fast speed, 3. in dual-task condition (while serially subtracting 7). Results are summarized in *Supplementary table 1*. At both sessions PSP patients performed significantly worse than HC regarding gait velocity and step length in all gait conditions. Cadence and step time were significantly reduced in PSP patients only in fast gait condition. The CoV of step length and step time were significantly increased in PSP at both sessions and in all conditions. Also, patients presented more gait asymmetry, especially in dual-task conditions. The only significant difference between PSP at baseline and follow-up was the step length in ‘preferred’ and ‘fast speed’ condition with worse performance at follow-up.

| gait parameter | gait condition | PSP at baseline | | PSP at follow-up | | HC | | *p*-values | | |
|---|---|---|---|---|---|---|---|---|---|---|
| | | | | | | | | PSP BL vs. HC | PSP FU vs. HC | PSP FU vs. BL |
| **velocity (cm/s)** | normal | 82.34 | ±19.33 | 75.94 | ±24.42 | 115.65 | ±23.26 | **0.001** | **0.002** | 0.268 |
| | fast | 111.48 | ±23.71 | 107.58 | ±27.46 | 203.53 | ±30.79 | **0.000** | **0.000** | 0.502 |
| | dual task | 57.39 | ±23.52 | 60.41 | ±27.07 | 94.37 | ±25.24 | **0.002** | **0.007** | 0.626 |
| **cadence (steps/min)** | normal | 97.58 | ±10.74 | 96.07 | ±14.90 | 101.03 | ±10.82 | 0.448 | 0.535 | 0.761 |
| | fast | 116.51 | ±14.58 | 117.06 | ±18.66 | 143.71 | ±13.80 | **0.000** | **0.001** | 0.773 |
| | dual task | 85.82 | ±20.34 | 87.23 | ±21.96 | 87.67 | ±5.35 | 0.730 | 0.836 | 0.626 |
| **step length (cm)** | normal | 50.28 | ±8.34 | 46.37 | ±10.70 | 68.16 | ±8.28 | **0.000** | **0.000** | **0.025** |
| | fast | 57.35 | ±9.36 | 54.66 | ±9.95 | 84.81 | ±8.41 | **0.000** | **0.000** | **0.042** |
| | dual task | 40.29 | ±12.16 | 39.81 | ±11.74 | 63.74 | ±9.33 | **0.000** | **0.000** | 0.855 |
| **step time (s)** | normal | 0.62 | ±0.08 | 0.64 | ±0.11 | 0.60 | ±0.07 | 0.435 | 0.535 | 0.749 |
| | fast | 0.53 | ±0.08 | 0.53 | ±0.09 | 0.42 | ±0.04 | **0.000** | **0.001** | 0.952 |
| | dual task | 0.88 | ±0.70 | 0.87 | ±0.56 | 0.71 | ±0.13 | 1.000 | 0.836 | 0.542 |
| **CoV step length (%)** | normal | 6.39 | ±2.68 | 8.35 | ±6.76 | 2.93 | ±1.06 | **0.000** | **0.001** | 0.268 |
| | fast | 5.84 | ±2.98 | 6.61 | ±4.00 | 3.26 | ±1.38 | **0.016** | **0.018** | 0.426 |
| | dual task | 14.14 | ±10.57 | 20.28 | ±29.25 | 4.20 | ±1.75 | **0.000** | **0.001** | 0.903 |
| **CoV step time (%)** | normal | 7.05 | ±2.99 | 7.59 | ±4.04 | 3.15 | ±0.74 | **0.000** | **0.000** | 0.391 |
| | fast | 7.61 | ±5.08 | 6.75 | ±3.82 | 3.80 | ±1.47 | **0.007** | **0.011** | 0.583 |
| | dual task | 18.88 | ±18.91 | 23.61 | ±33.88 | 4.84 | ±2.46 | **0.001** | **0.002** | 1.000 |
| **asymmetry index** | normal | 1.08 | ±0.06 | 1.09 | ±0.09 | 1.02 | ±0.02 | **0.003** | **0.016** | 1.000 |
| | fast | 1.09 | ±0.07 | 1.07 | ±0.06 | 1.03 | ±0.02 | **0.004** | 0.063 | 0.153 |
| | dual task | 1.12 | ±0.13 | 1.11 | ±0.12 | 1.03 | ±0.02 | **0.012** | **0.014** | 0.855 |

**Supplementary table 1 | Gait characteristics for different gait conditions across groups.**
*p*-values are obtained from non-parameteric tests ($n$ = 14). Bold font indicates significant *p*-value.
